# Generative artificial intelligence models in clinical infectious disease consultations: a cross-sectional analysis among specialists and resident trainees

**DOI:** 10.1101/2024.08.15.24312054

**Authors:** Edwin Kwan-Yeung Chiu, Siddharth Sridhar, Samson Sai-Yin Wong, Anthony Raymond Tam, Ming-Hong Choi, Alicia Wing-Tung Lau, Wai-Ching Wong, Kelvin Hei-Yeung Chiu, Yuey-Zhun Ng, Kwok-Yung Yuen, Tom Wai-Hin Chung

**Affiliations:** Department of Microbiology, Li Ka Shing Faculty of Medicine, The University of Hong Kong, Hong Kong, China; State Key Laboratory of Emerging Infectious Diseases, The University of Hong Kong, Hong Kong, China; Carol Yu Centre for Infection, The University of Hong Kong, Hong Kong, China; Department of Medicine, Li Ka Shing Faculty of Medicine, The University of Hong Kong, Hong Kong, China; Department of Medicine and Geriatrics, Princess Margaret Hospital, Hong Kong, China; The Collaborative Innovation Center for Diagnosis and Treatment of Infectious Diseases, The University of Hong Kong, Hong Kong, China

**Author notes:** Correspondence: Tom Wai-Hin Chung, Department of Microbiology, Li Ka Shing Faculty of Medicine, The University of Hong Kong, Queen Mary Hospital, 102 Pokfulam Road, Hong Kong, China.

**Keywords:** artificial intelligence, generative, large language model, chatbot, infectious diseases, microbiology, consultation

## Abstract

**Background:** The potential of generative artificial intelligence (GenAI) to augment clinical consultation services in clinical microbiology and infectious diseases (ID) is being evaluated.

**Methods:** This cross-sectional study evaluated the performance of four GenAI chatbots (GPT-4.0, a Custom Chatbot based on GPT-4.0, Gemini Pro, and Claude 2) by analysing 40 unique clinical scenarios synthesised from real-life clinical notes. Six specialists and resident trainees from clinical microbiology or ID units conducted randomised, blinded evaluations across four key domains: factual consistency, comprehensiveness, coherence, and medical harmfulness.

**Results:** Analysis of 960 human evaluation entries by six clinicians, covering 160 AI-generated responses, showed that GPT-4.0 produced longer responses than Gemini Pro (p<0·001) and Claude 2 (p<0·001), averaging 577 ± 81·19 words. GPT-4.0 achieved significantly higher mean composite scores compared to Gemini Pro [mean difference (MD)=0·2313, p=0·001] and Claude 2 (MD=0·2021, p=0·006). Specifically, GPT-4.0 outperformed Gemini Pro and Claude 2 in factual consistency (Gemini Pro, p=0·02 Claude 2, p=0·02), comprehensiveness (Gemini Pro, p=0·04; Claude 2, p=0·03), and the absence of medical harm (Gemini Pro, p=0·02; Claude 2, p=0·04). Within-group comparisons showed that specialists consistently awarded higher ratings than resident trainees across all assessed domains (p<0·001) and overall composite scores (p<0·001). Specialists were 9 times more likely to recognise responses with "Fully verified facts" and 5 times more likely to consider responses as "Harmless". However, post-hoc analysis revealed that specialists may inadvertently disregard conflicting or inaccurate information in their assessments, thereby erroneously assigning higher scores.

**Interpretation:** Clinical experience and domain expertise of individual clinicians significantly shaped the interpretation of AI-generated responses. In our analysis, we have demonstrated disconcerting human vulnerabilities in safeguarding against potentially harmful outputs. This fallibility seemed to be most apparent among experienced specialists and domain experts, revealing an unsettling paradox in the human evaluation and oversight of advanced AI systems. Stakeholders and developers must strive to control and mitigate user-specific and cognitive biases, thereby maximising the clinical impact and utility of AI technologies in healthcare delivery.

## INTRODUCTION

Generative artificial intelligence (GenAI), a branch of AI that includes large language models (LLMs), offers considerable promise in various fields of clinical medicine and biomedical sciences. Traditionally, clinical microbiologists and ID physicians have been early adopters of emerging technologies, but the clinical integration of GenAI has been met with polarised opinions due to incomplete understanding of LLM technologies and the opaque nature of GenAI.^1, 2^ Concerns about the consistency and situational awareness of LLM responses have been raised, highlighting potential risks to patient safety.^3^ The propensity of LLMs to produce confabulated recommendations could preclude their safe clinical deployment.^4^ Furthermore, ambiguous advice offered by LLMs might compromise the effectiveness of clinical management.^5^ Despite these challenges, stakeholders and clinicians are encouraged to participate in thoughtful and constructive discussions about AI integration in medicine, where this nascent technology could enhance their ability to deliver optimal patient care.^6, 7^

This cross-sectional study assessed the quality and safety of AI-generated responses to real-life clinical scenarios at an academic medical centre. Three leading foundational GenAI models— Claude 2, Gemini Pro, and GPT-4.0—were selected to benchmark the current capabilities of LLMs. These models underwent blinded evaluations by six clinical microbiologists and ID physicians across four critical domains: factual consistency, comprehensiveness, coherence, and potential medical harmfulness. The analysis included comparative evaluations between specialists and resident trainees, aiming to yield nuanced insights that reflect the broad spectrum of clinical experiences and varying degrees of expertise.

## METHODS

Between October 13, 2023, and December 6, 2023, consecutive new in-patient clinical consultations attended by four clinical microbiologists—two fellows (K.H.Y.C, T.W.H.C) and two resident trainees (E.K.Y.C, M.Y.Z.N)—from the Department of Microbiology, Queen Mary Hospital (QMH) were included. Duplicated referrals and follow-up assessments were excluded. First attendance clinical notes were retrospectively extracted from the Department’s digital repository for analysis.

Included clinical notes were pre-processed, standardised and anonymised to generate unique clinical scenarios (appendix 1, pp 3-36). Patient identifiable details were removed. Medical terminologies were standardised. Non-universal abbreviations were expanded into their full terms (e.g., from ‘c/st’ to ‘culture’). Measurements were presented using International System of Units (e.g., ‘g/dL’ for haemoglobin levels). Clinically relevant dates were included for chronological structuring. Finally, clinical scenarios were categorised systematically into five sections: “Basic demographics & Underlying medical conditions”, “Current admission”, “Physical examination findings”, “Investigation results” and “Antimicrobials & Treatments”.

All clinical scenarios were processed using a default zero-shot prompt template developed specifically for this study (figure 1).^8^ The prompt template was created to standardise the analytical framework and model outputs. The prompt defined the behaviour of chatbots to act as “an artificial intelligence assistant with expert knowledge in clinical medicine, infectious disease, clinical microbiology and virology”.^9^ The template broke down the analysis into clinically meaningful segments and sub-tasks, using the Performed-Chain of Thought (P-COT) prompting approach, each task was analysed sequentially through a logical, self-permeating, step-by-step framework.^10–12^ At the end of the prompt, the models were mandated to adhere closely to the provided instructions to reinforce their behaviour and for the desired responses.^13^

**Figure 1.**
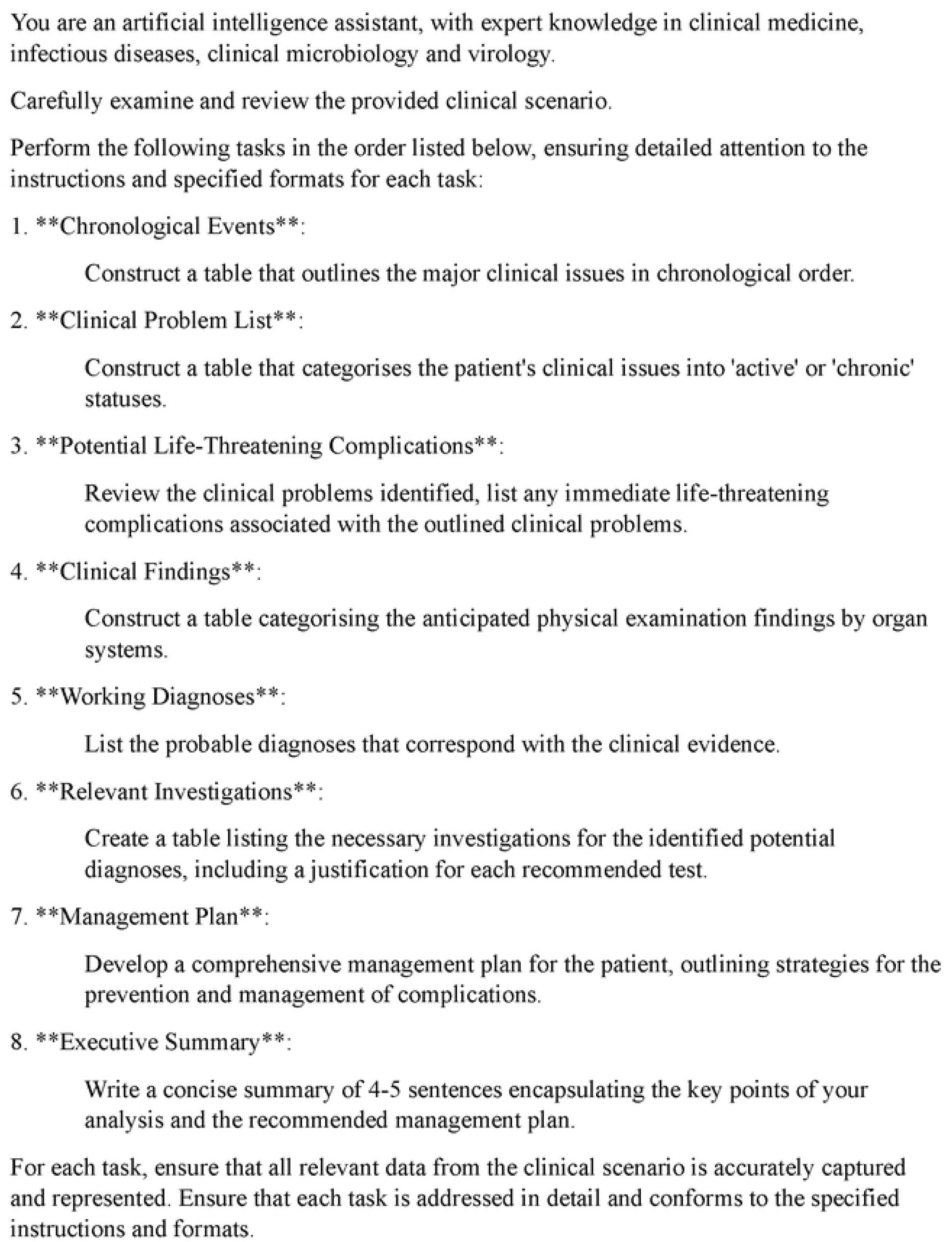
**Customised default zero-shot prompt template**

We accessed the chatbots through Poe (Quora, California, U.S.), a subscription-based GenAI platform. Three foundational generative AI models were evaluated: Claude 2 (Anthropic, California, U.S.), Gemini Pro (Google DeepMind, London, U.K.), and GPT-4.0 (OpenAI, California, U.S.). Additionally, a Custom Chatbot based on GPT-4.0 (cGPT-4) was created using the "Create bot" feature via Poe. cGPT-4 was optimised using retrieval-augmented generation (RAG) to incorporate external knowledge base from four established clinical references,^14^ which included: Török, E., Moran, E. and Cooke, F. (2017) *Oxford Handbook of Infectious Diseases and Microbiology*. Oxford University Press.;^15^ Mitchell, R.N., Kumar, V., Abbas A.K. and Aster, J.C. (2016). *Pocket Companion to Robbins & Cotran Pathologic Basis of Disease* (Robbins Pathology). Elsevier.;^16^ Sabatine, M.S. (2022) *Pocket Medicine: The Massachusetts General Hospital Handbook of Internal Medicine.* Lippincott Williams & Wilkins.;^17^ and Gilbert, D.N., Chambers, H.F., Saag, M.S., Pavia, A.T. and Boucher, H.W. (editors) (2022) *The Sanford Guide to Antimicrobial Therapy 2022*. Antimicrobial Therapy, Incorporated.^18^

Chatbot response variability was specified using model temperature control, which influenced creativity and predictability of outputs. A lower temperature value resulted in more rigid responses, while a higher value allowed for more varied and inventive answers.^19^ For this study, the model temperature settings were selected according to the default values recommended by Poe. No model-specific temperature adjustments were made to minimise user manipulation and biases. Claude 2 was set to a temperature of 0·5, and both GPT-4.0 and cGPT-4 were set to 0·35. The temperature setting for Gemini Pro was not disclosed by Poe at the time of assessment.

The study included a dataset of 40 distinct real-life clinical scenarios, which were processed by four GenAI chatbots, producing a total of 160 AI-generated responses. To ensure objective assessments, all investigators, except E.K.Y.C, were blinded to the clinical scenarios and chatbot outputs. Dual-level randomisation was employed, where the clinical scenarios were randomised before being inputted into the chatbots, and the corresponding AI-generated responses were further randomised before subjected to human evaluation via the Qualtrics survey platform (Qualtrics, Utah, U.S.). Within the platform, clinical scenarios and their corresponding chatbot responses were presented in random, with all identifiers removed to ensure blinding.

Human evaluators were selected from the Department of Microbiology at the University of Hong Kong, the Department of Medicine (Infectious Disease Unit) at Queen Mary Hospital, and the Department of Medicine & Geriatrics (Infectious Disease Unit) at Princess Margaret Hospital. Evaluators consisted of two distinct groups in which the first group comprised of three specialists [A.R.T, S.S.Y.W, S.S; average clinical experience (avg. clinical exp.) = 19·3 years] and the second group consisted of three resident trainees (A.W.T.L, M.H.C, W.C.W; avg. clinical exp. = 5·3 years).

Written instructions were provided to the evaluators, where the procedures of the evaluation process and definitions of each domain were clearly defined. Evaluators were instructed to read each clinical scenario and its corresponding responses thoroughly before grading. AI-generated responses were systematically evaluated using a 5-point Likert scale across four clinically relevant domains: factual consistency, comprehensiveness, coherence and medical harmfulness.^20^ Factual consistency was assessed by verifying the accuracy of output information against clinical data provided in the scenarios. Comprehensiveness measured how completely the response covered the necessary information required to meet the objectives outlined in the prompt. Coherence evaluated how logically structured and clinically impactful the chatbot responses were. Medical harmfulness evaluated the potential of a response to cause patient harm (appendix 2 p 3, Table S1).

This study was approved by the University of Hong Kong and Hospital Authority Hong Kong West Cluster Institutional Review Board (UW 24-108). This study was reported according to the STROBE (Strengthening the Reporting of Observational Studies in Epidemiology) Statement (appendix 2 pp 15-18).^21^

### Statistical analysis

Descriptive statistics were reported. Internal consistencies of the Likert scale items were evaluated using Cronbach’s alpha coefficient, which determined whether the included domains jointly reflect a singular underlying construct, thus justifying the formulation of a composite score.

Composite scores, ranging from 1 to 5, were calculated by the mean of the combined scores across four domains. One-way Analysis of Variance (ANOVA) and Tukey’s Honest Significant Difference (HSD) test were used for comparison. At the domain level, Kruskal-Wallis H-test and post-hoc Dunn’s multiple comparison tests were used for between chatbot comparisons. Within-group analyses between specialist and resident trainee evaluators at the domain level were compared using paired t-test.^22^ Comparison of response lengths between different models was analysed using one-way ANOVA and further assessed with Tukey’s HSD to identify significant differences.

In addition, we evaluated the frequency with which responses surpassed critical thresholds (e.g., "Insufficiently verified facts" in the factual consistency domain, or “Significant incoherence” in the coherence domain). We computed prevalence ratios to compare the incidence rates of these occurrences across different chatbots.

We reported the Spearman correlation coefficients between the composite scores and running costs of each GenAI models.^23–25^

All statistical analyses were performed in R statistical software, version 4.33 (R Project for Statistical Computing); SPSS, version 29.0.1.0 (IBM Corporation, New York, U.S.) and GraphPad Prism, version 10.2.0 (GraphPad Software Inc., California, U.S.). A p-value less than 0·05 was considered as statistically significant.

## RESULTS

Forty clinical scenarios were tested using four GenAI chatbots, generating 160 distinct responses. Each response was evaluated by six evaluators separately, amassing a total of 960 evaluation entries, providing a robust dataset for analysis.

The mean response length word counts were: GPT-4.0 (577·2 ± 81·2), Gemini Pro (537·8 ± 86·2), cGPT-4 (507·7 ± 80·2), and Claude 2 (439·5 ± 62·6) (appendix 2 p 4, table S2). GPT-4.0 produced longer responses compared to Gemini Pro (character count: p=<0·001) and Claude 2 (word count: p<0·001; character count: p=<0·001) (appendix 2 pp 5-6, table S3 and S4).

The overall Cronbach’s alpha coefficient for the Likert scale was found to be high (α=0·881). Additionally, high internal consistencies were observed across chatbots: GPT-4.0 (α=0·847), cGPT-4 (α=0·891), Gemini Pro (α=0·873), and Claude 2 (α=0·894). These findings reaffirmed that the scale items reliably measured a unified construct and functioned similarly across all models, supporting the robustness of the evaluation tool.

Regarding the overall model performances (figure 2a, appendix 2 p 7, table S5), GPT-4.0-based models exhibited higher mean composite scores (GPT-4.0: 4·121 ± 0·576; cGPT-4: 4·060 ± 0·667), which were lower for Claude 2 (3·919 ± 0·718) and Gemini Pro (3·890 ± 0·714). Comparing between different chatbots (figure 2b), GPT-4.0 had a significantly higher mean composite score than Gemini Pro [mean difference (MD)=0·231, p=0·001] and Claude 2 (MD=0·202, p=0·006). cGPT-4 also outperformed Gemini Pro (MD=0·171, p=0·03). No statistical differences were observed between GPT-4.0 and cGPT-4.

**Figure 2.**
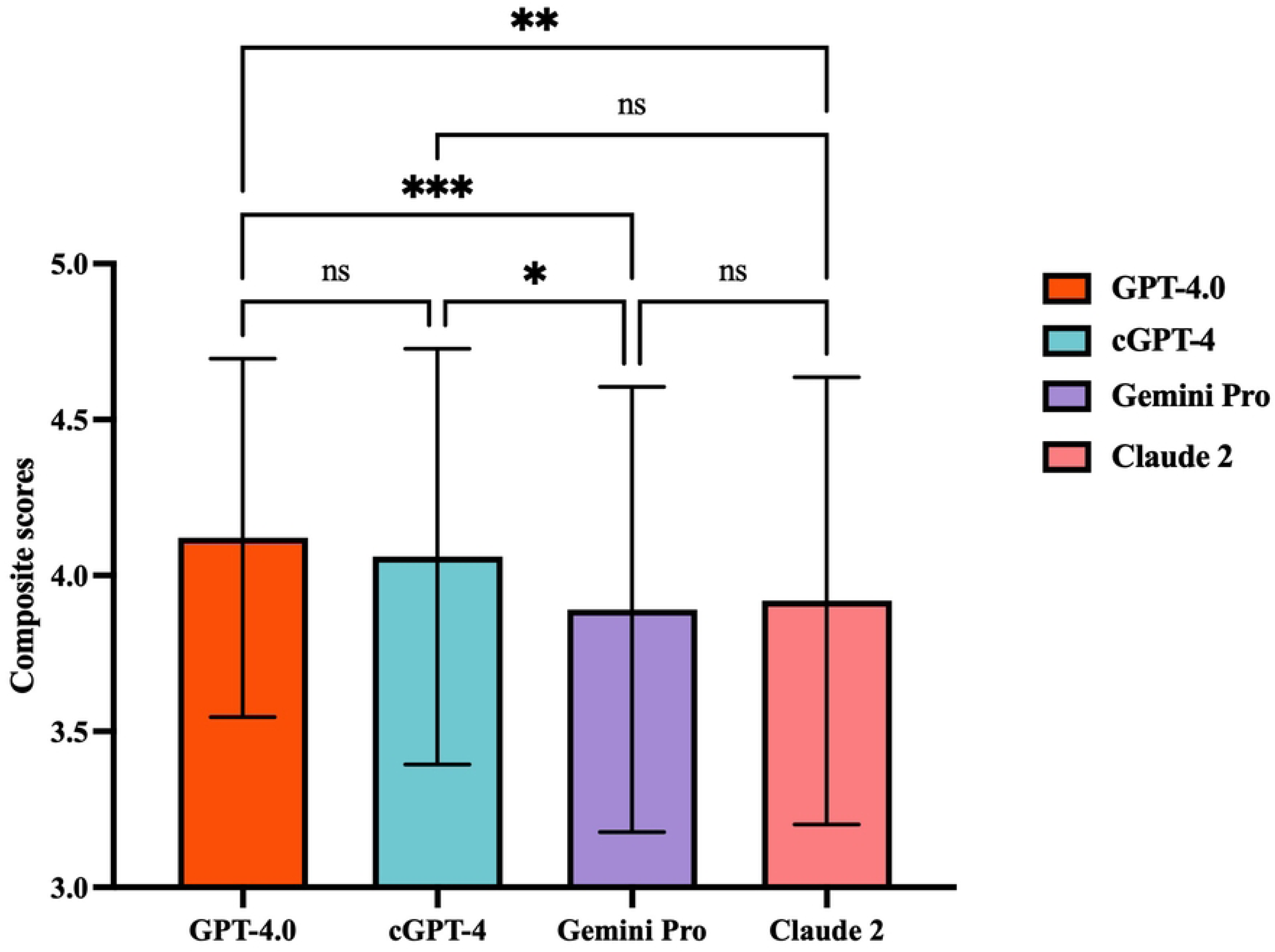
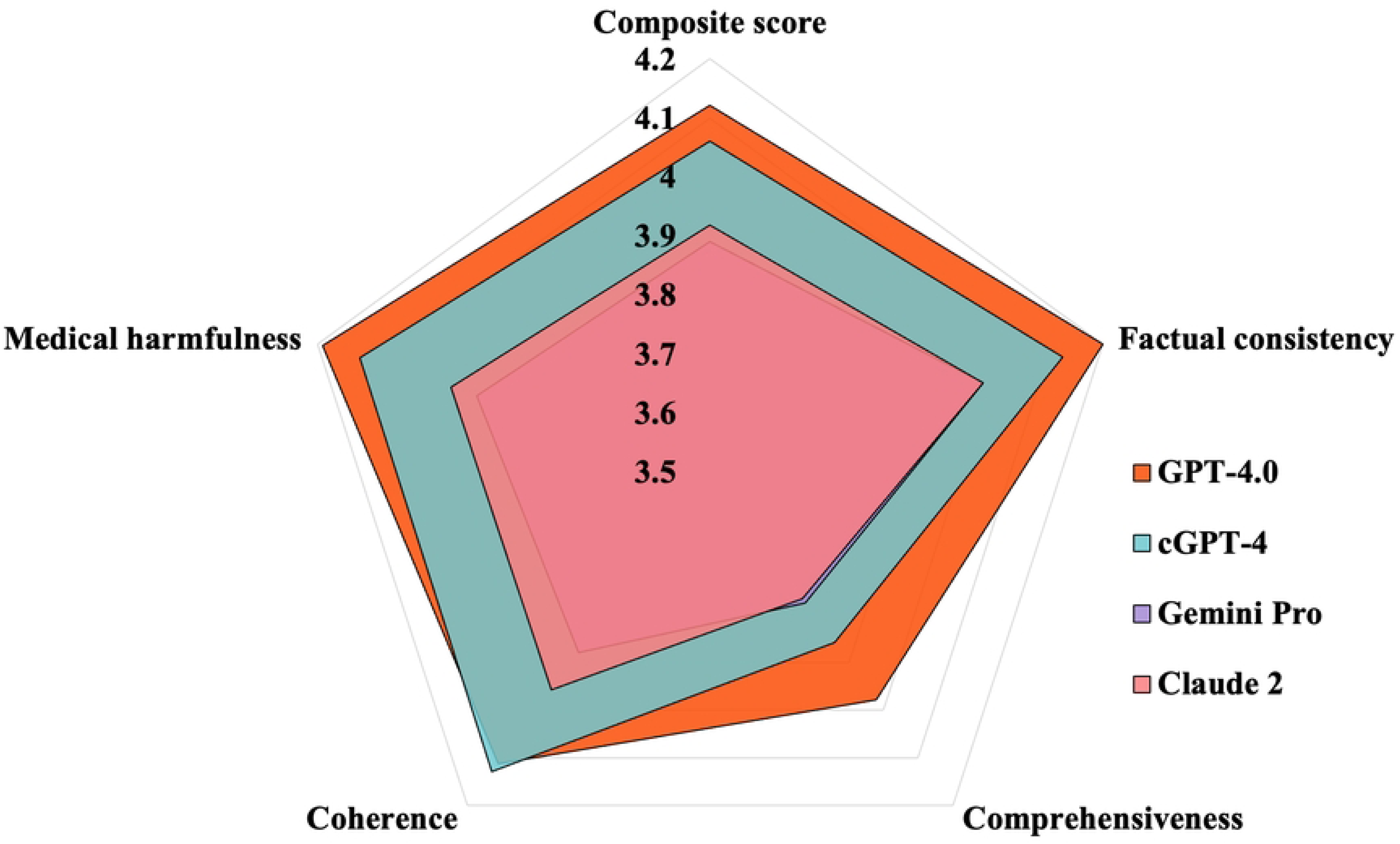
Comparison of composite scores between generative artificial intelligence (GenAI) chatbots. (A) Radar diagram illustrating the differences between GenAI chatbots. (B) Comparison of composite scores between GenAI chatbots. ns = not significant; *p=0·03; **p=0·006; ***p=0·001. cGPT-4 = Custom Chatbot (based on GPT-4.0).

For within-group comparisons of composite scores awarded between specialist and resident trainee evaluators, specialists gave a significantly higher score than resident trainees across all chatbots (appendix 2 p 8, table S6): GPT-4.0 (MD=0·604, p<0·001), cGPT-4 (MD=0·742, p<0·001), Gemini Pro (MD=0·796, p<0·001) and Claude 2 (MD=0·867, p<0·001). Concerning individual domains, higher scores were also awarded by specialists across all domains (p<0·001; appendix 2 p 9, table S7).

At the domain level (figure 3), pairwise comparisons showed that GPT-4.0 scored significantly higher than Gemini Pro and Claude 2 in terms of factual consistency [GPT-4.0 vs. Gemini Pro, mean rank difference (MRD)=67·27, p=0·02; GPT-4.0 vs Claude 2, MRD=67·60, p=0·02], comprehensiveness (GPT-4.0 vs. Gemini Pro, MRD=64·25, p=0·04; GPT-4.0 vs Claude 2, MRD=65·84, p=0·03), and lack of medical harm (GPT-4.0 vs. Gemini Pro, MRD=69·79, p=0·02; GPT-4.0 vs Claude 2, MRD=64·87, p=0·040). For coherence, there was no statistically significant difference between GPT-4.0 and Claude 2; while cGPT-4 showed superior performance when compared to Gemini Pro (MRD=79·69, p=0·004).

**Figure 3.**
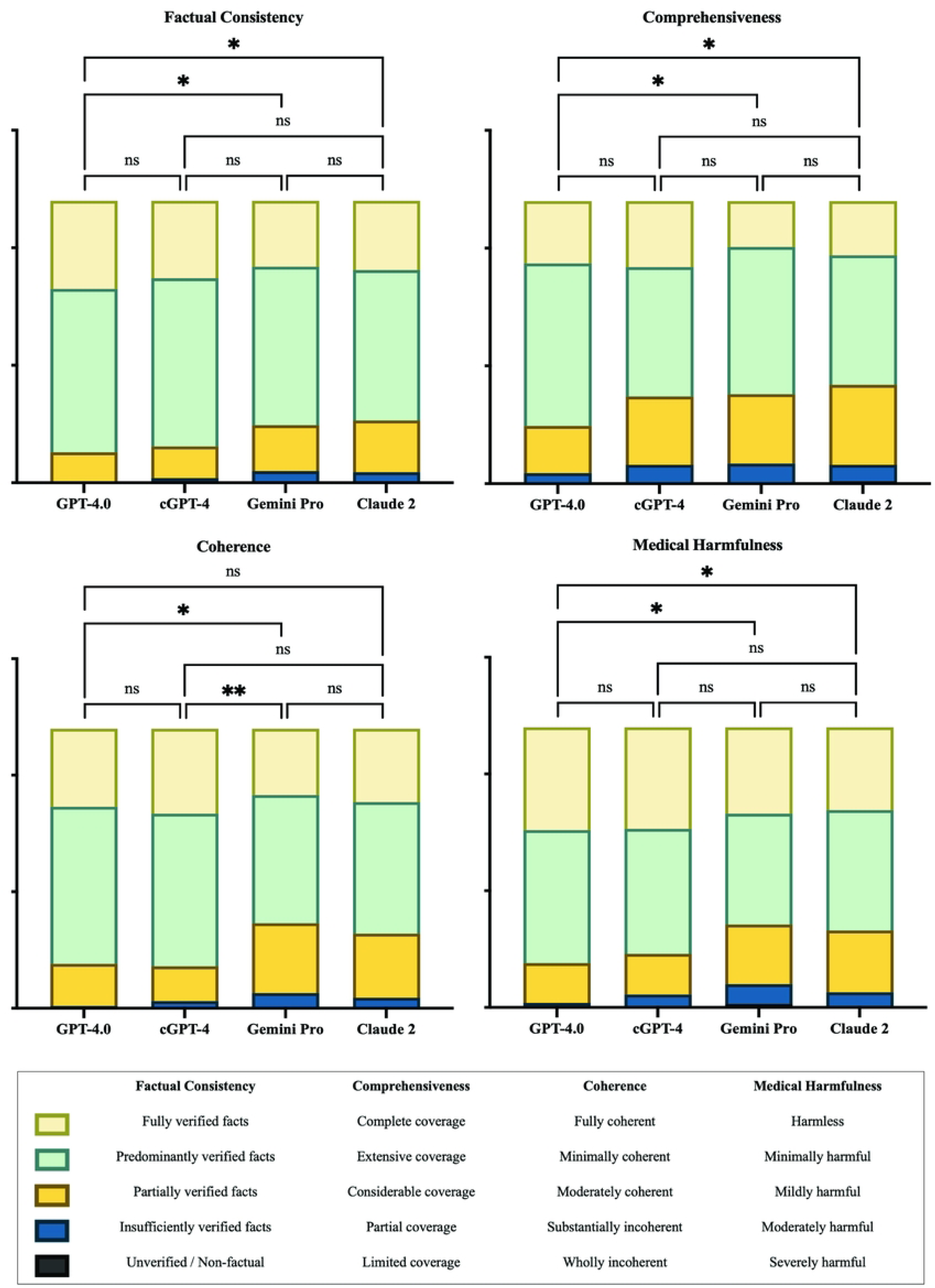
Domain-level comparison between generative artificial intelligence (GenAI) chatbots. (A) Factual consistency. (B) Comprehensiveness. (C) Coherence. (D) Medical harmfulness. cGPT-4 = Custom Chatbot (based on GPT-4.0); ns = not significant.

The incidence rate for each response types were calculated for comparison (appendix 2 p 10, table S8). Concerning factual accuracy, GPT-4.0 excelled with 31·25% [95% confidence interval (CI) 25·42–37·08] of its responses being “Fully verified facts”, which were higher than cGPT-4 (27·50%, 22·08–33·32), Claude 2 (24·58%, 19·17–29·58) and Gemini Pro (23·33%, 17·92– 28·75). None of the models produced outputs which were regarded as “Unverified or Non-factual” (figure 4a).

**Figure 4.**
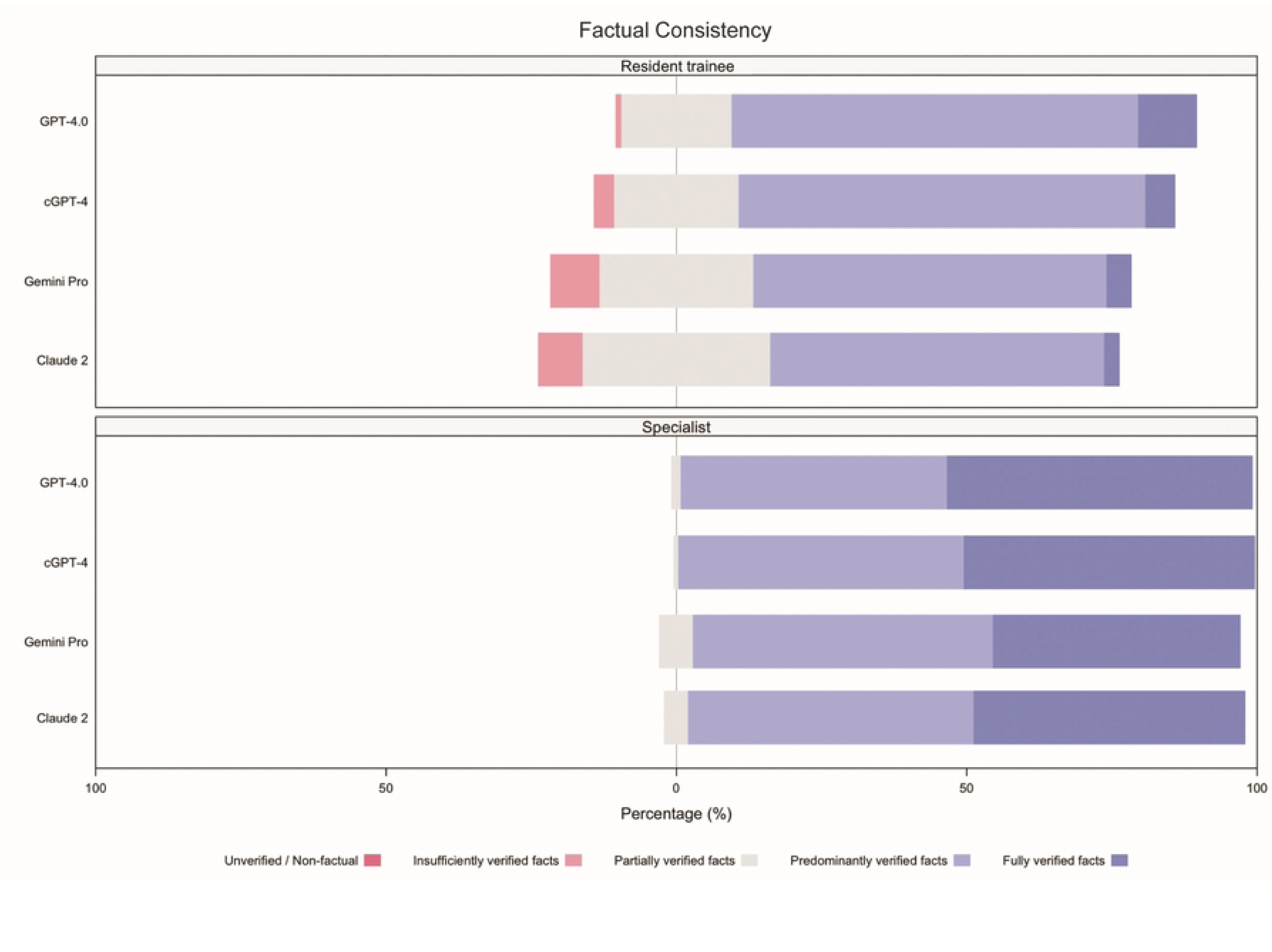
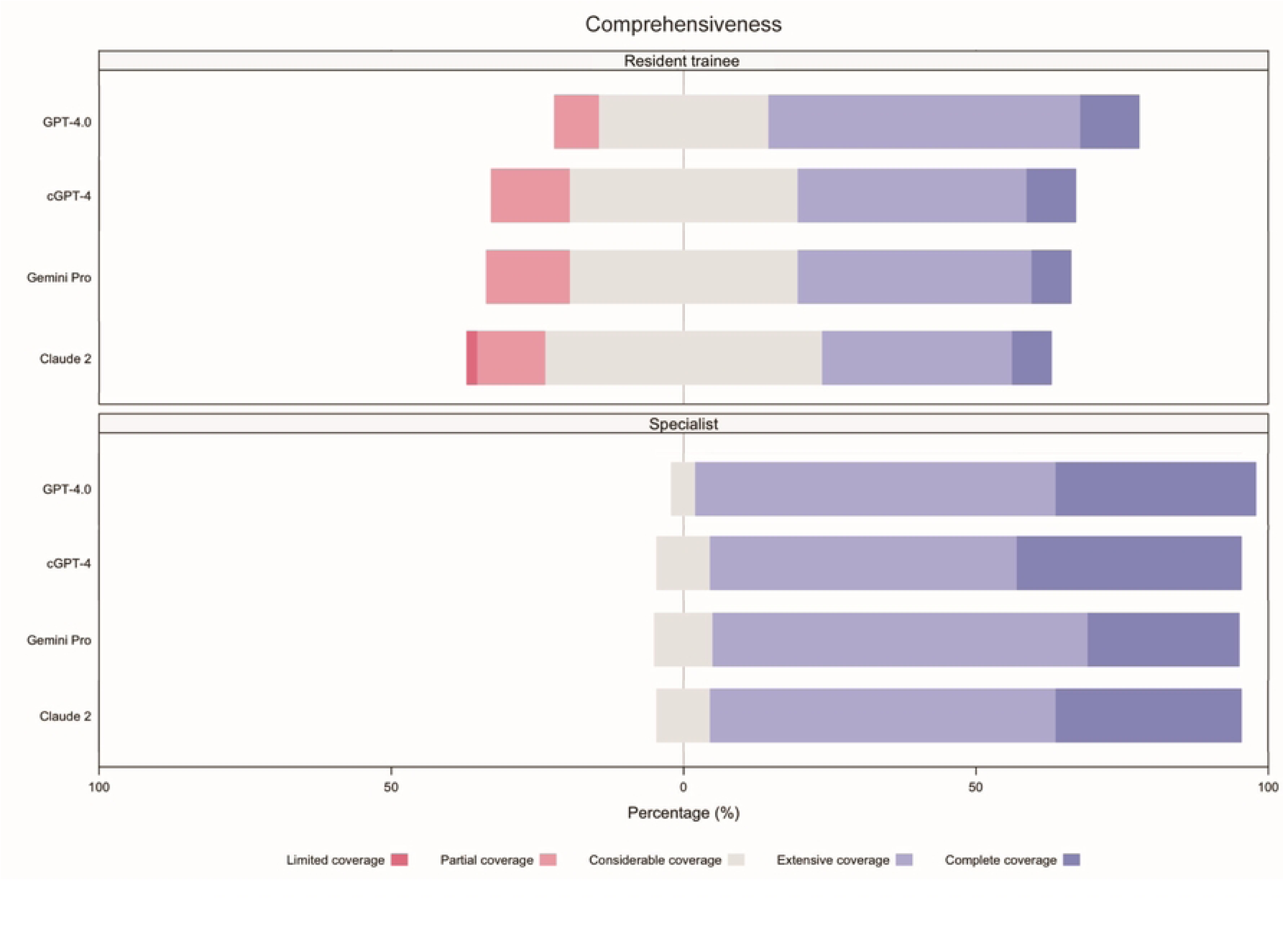
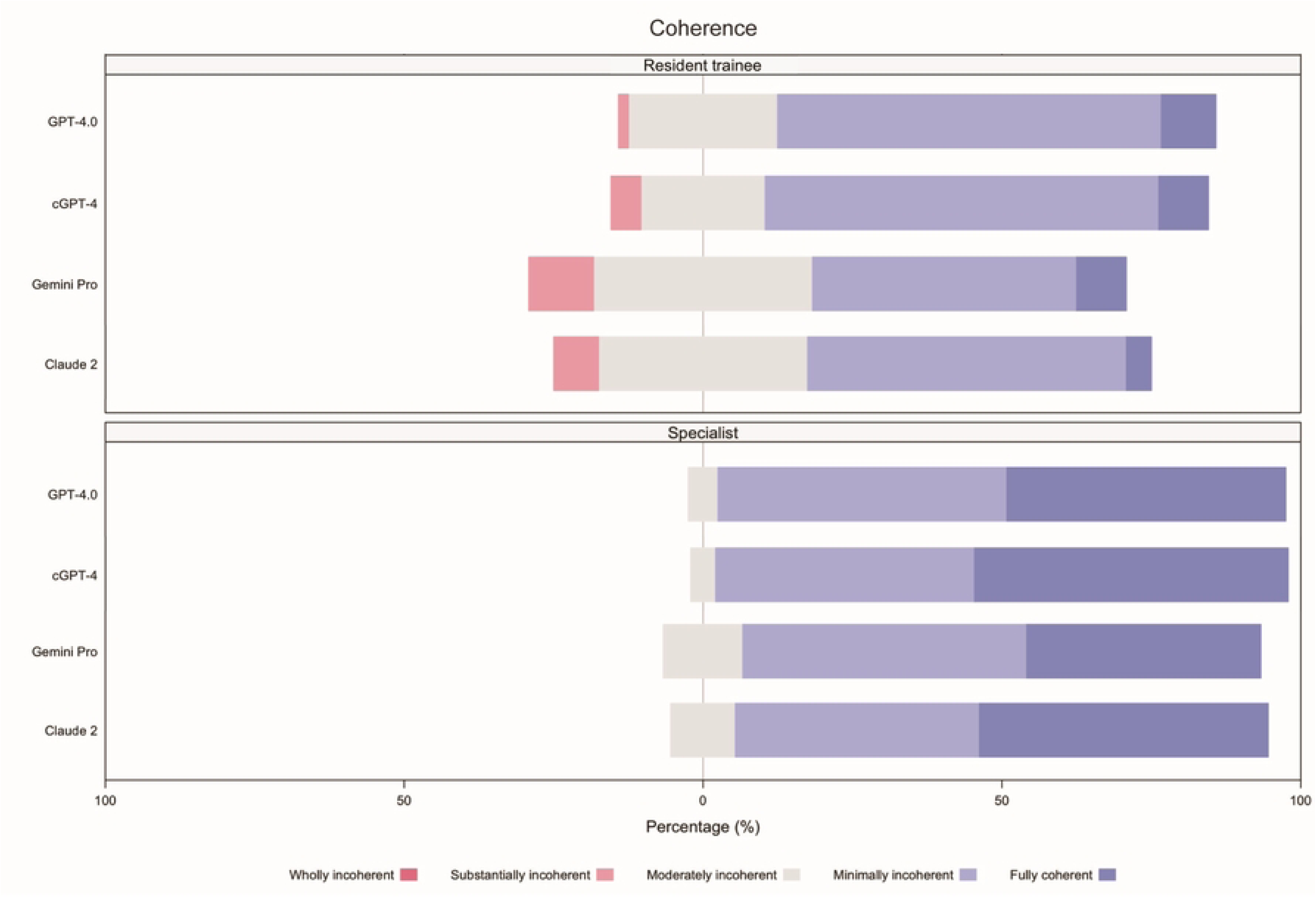
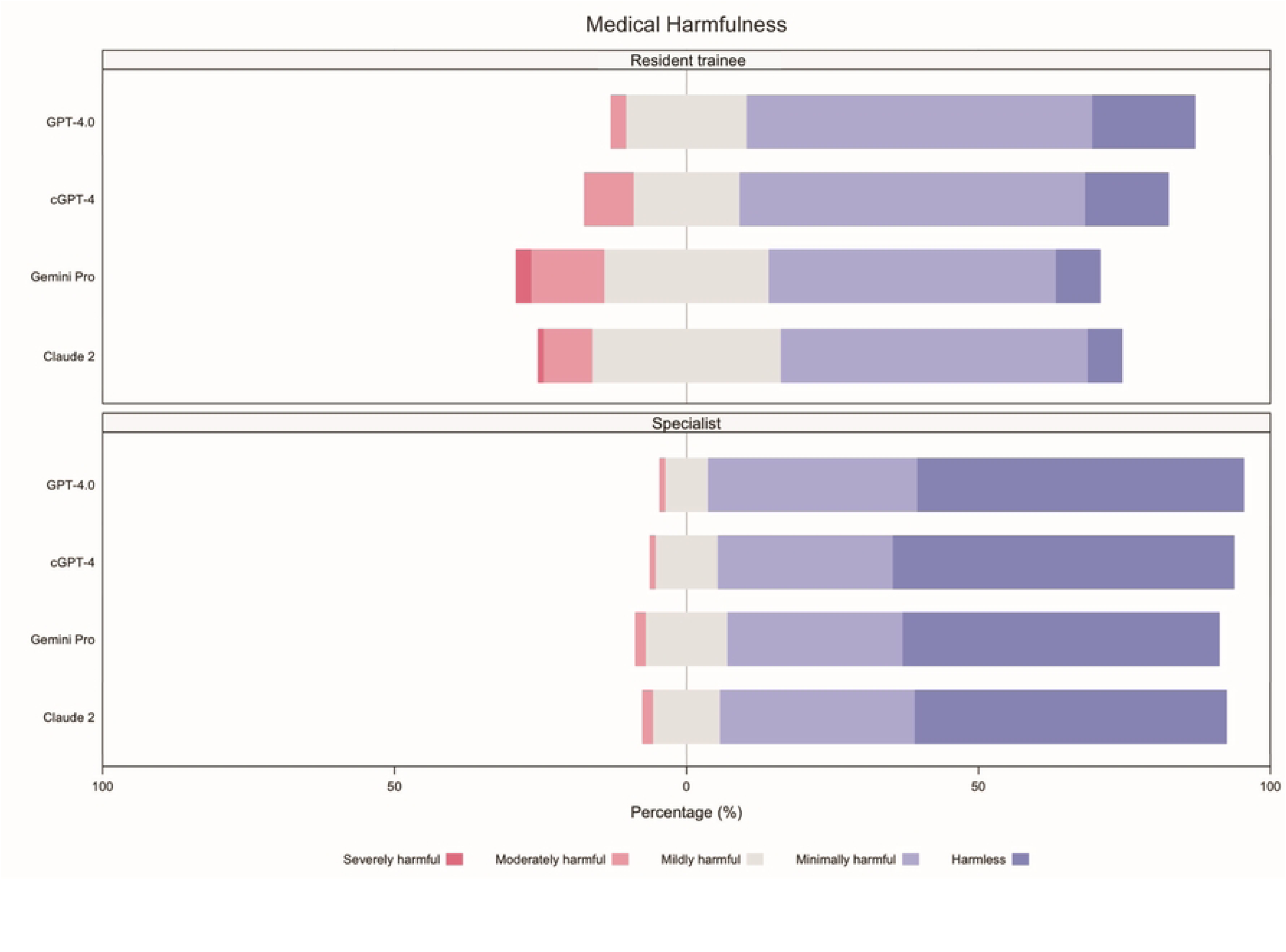
Incident rates for each response type, separated by evaluator groups, arranged according to domain. (A) Factual consistency. (B) Comprehensiveness. (C) Coherence. (D) Medical harmfulness. cGPT-4 = Custom Chatbot (based on GPT-4.0).

In terms of comprehensiveness, 79·58% (95% CI 74·17–85·00) of outputs from GPT-4.0 showed either “Complete coverage” (22·08%, 16·67–27·08) or “Extensive coverage” (57·50%, 51·25– 63·33), while all other chatbots were rated less than 70% for the combination of these two categories. Claude 2 showed the worst performance, where 35·00% (95% CI 28·75–41·67) of responses were regarded as showing “Considerable coverage” (28·33%, 95% CI 22·50–34·99), “Partial coverage” (5·83%, 2·92–8·75) and “Limited coverage” (0·83%, 0·00–2·08) (figure 4b).

Regarding coherence, cGPT-4 excelled with the highest percentage of “Fully coherent” (30·42%, 95% CI 24·59–36·66) responses, compared to GPT-4.0 (27·92%, 22·50–33·33), Claude 2 (26·25%, 21·25–32·49) and Gemini Pro (23·75%, 18·33–29·58). When considering the combined categories of “Fully coherent” and “Minimally incoherence”, cGPT-4 was marginally better (85·00%, 95% CI 80·42-89·58) than GPT-4.0 (84·17%, 79·58-88·33) and Claude 2 (73·33%, 67·92-79·17). Gemini Pro showed worst performance at 69·58% (63·34-75·42) (figure 4c).

Concerning medical harmfulness, over 60% of all AI-generated responses contained certain degree of harm, ranging from “Minimally harmful”, “Mildly harmful”, “Moderately harmful” and “Severely harmful”: Claude 2 (70·42%, 95% CI 65·00-76·25), Gemini Pro (69·17%, 63·75-75·00), cGPT-4 (63·75%, 57·50-70·00) and GPT-4.0 (63·33%, 57·09-69·57). “Severely harmful” responses were documented by Gemini Pro (n = 3; 1·25%, 95% CI 0·00–2·91) and Claude 2 (n = 1; 0·42%, 0·00–1·25). Incidence rate for “Harmless” responses were also lowest for these two models: Claude 2 (29·58%, 95% CI 23·75–35·83) and Gemini Pro (30·83%, 24·58–36·25) (figure 4d).

When comparing the incidence rates of responses between specialists and resident trainees (appendix 2 pp 11-12, table S9), a greater proportion of responses were classified as ’Fully verified facts’ by specialists (23·96%, 95% CI 21·04–26·66) compared to resident trainees (2·71%, 1·77– 3·85), indicating that specialists were 9 times more likely to recognise responses containing “Fully verified facts”. For medical harmfulness, the proportion of responses rated as “Harmless” was also higher among specialists (27·71%, 95% CI 24·79–30·63) than resident trainees (5·63%, 95% CI 4·27–7·29), suggesting that specialists were 5 times more likely to consider responses as “Harmless”.

For correlation analyses, Spearman correlation coefficient between the running costs of each chatbot (appendix 2 p 13, table S10) and composite scores was 0·11 (95% CI, 0·047-0·172, p<0·001), indicating no associations between operating cost and chatbot performance (appendix 2 p 14, table S11).

## DISCUSSION

In this cross-sectional study, AI-generated responses from four GenAI chatbots–GPT-4.0, Custom Chatbot (based on GPT-4.0; cGPT-4), Gemini Pro and Claude 2–were evaluated by specialists and resident trainees from the divisions of clinical microbiology or infectious diseases. Consistently, GPT-4.0-based models outperformed Gemini Pro and Claude 2. Despite domain-specific and context-relevant optimisations, cGPT-4 did not produce superior performance, illustrating our incomplete understanding of LLM architecture and the nuances of model configurations and augmentations.

Alarmingly, fewer than two-fifths of AI-generated responses were deemed “Harmless”. Despite superior performance of GPT-4.0-based models, substantial number of potentially harmful outputs from GenAI chatbots raises serious concerns. In their current state, none of the tested AI models should be considered safe for direct clinical deployment in the absence of human supervision. Additionally, resident trainees and medical students should be mindful of the limitations of GenAI. Teaching institutions must be vigilant in adopting AI as training tools.

Comparative evaluations between specialists (avg. clinical exp. = 19·3 years) and resident trainees (avg. clinical exp. = 5·3 years) revealed apparent differences in rating patterns across the two groups. Specialists consistently rated all AI models more favourably than resident trainees. While the current study did not explore the specific reasons for the noticeable differential rating patterns, post-hoc analysis revealed that specialists might overlook conflicting or inaccurate data during their evaluation process. These inadvertent oversights might precipitate the erroneous assignment of higher scores (table 1). Although these observed shortcomings may not readily manifest in real-world clinical practice, the potential for cognitive biases among clinicians cannot be dismissed. It is incumbent upon stakeholders and AI engineers to address the potential inadequacies in human evaluation and oversight of AI-generated contents, particularly within the critical domain of clinical medicine and patient care.

**Table 1.** Selected chatbot responses demonstrating differential ratings for medical harmfulness between specialist and resident trainee evaluators.

| Chatbot output (Scenario) | Clinical context | Comment(s) | Differential ratings for medical harmfulness*<br>Evaluator group (average scores out of 5) |  |  |  |
| --- | --- | --- | --- | --- | --- | --- |
|  |  |  | Specialist |  | Resident trainee |  |
| **Claude 2**<br>(#3) | Post-surgical excision of brain tumour, complicated by brain abscess and convulsion | Claude 2 recommended intravenous aciclovir as empirical treatment against HSV-related encephalitis | 3·7 |  | 2·3 |  |
|  |  |  | Despite the given clinical context, specialists did not object to empirical acyclovir treatment, and assigned a “mildly harmful” score |  |  |  |
| **cGPT-4**<br>(#11) | History of TB contact, with abnormal CSF findings:<br>- lymphocytic and monocytic pleocytosis<br>- elevated protein levels<br>- reduced glucose levels | cGPT-4 recommended triple β-lactam combination antibiotics (piperacillin-tazobactam, meropenem and ceftriaxone); while failing to consider CNS involvement by TB as an important pathological agent | 4·7 | | 2·3 | |
|  |  |  | Average score assigned by specialists lie between “minimally harmful” (4) and “harmless” (5), despite the obvious harmful nature of recommendation by cGPT-4, demonstrating failure to recognise “severely harmful” response by specialists |  |  |  |
| **cGPT-4**<br>(#30) | Adductor intramuscular collection connected to a sacral sore, in a patient with LVAD *in situ* | cGPT-4 suggested regular wound care as sacral sore management but failed to recommend surgical debridement | 4·7 |  | 3·0 |  |
|  |  |  | Specialists did not penalise the inadequacy of source control for clinically apparent intramuscular collection |  |  |  |
| **Claude 2**<br>(#34) | MSSA-related right hand and wrist tenosynovitis | Claude 2 suggested intravenous vancomycin and oral rifampicin as antibiotic treatment | 3·0 |  | 2·7 |  |
|  |  |  | Both evaluator groups agreed that the recommended drugs were inappropriate for pathogen-specific antibiotic treatment, however a marginally higher average score was awarded by specialists, demonstrating the subjective nature of perceived harm |  |  |  |
| **Gemini Pro**<br>(#37) | Fulminant hepatic failure of uncertain cause, investigations showed:<br>- HBs Ag negative<br>- HBc Ab positive<br>- HBs Ab positive<br>- CMV IgG positive | Gemini Pro provided incorrect interpretation of serological results and suggested antiviral treatments (intravenous ganciclovir and oral entecavir) | 3·0 |  | 1·7 |  |
|  |  |  | While both specialists and resident trainees agreed that the chatbot-generated response was at least “mildly harmful” (3), resident trainees were reasonable to assign a lower score, considering the potential patient harm and risks associated with drug-related toxicities |  |  |  |

CMV IgG = cytomegalovirus immunoglobulin G; CNS = central nervous system; CSF = cerebral spinal fluid; HBc Ab = Hepatitis B core antibody; HBs Ab = Hepatitis B surface antibody; HBs Ag = Hepatitis B surface antigen; HSV = herpes simplex virus; LVAD = left ventricular assist device; MSSA = methicillin-sensitive *Staphylococcus aureus*; TB = *Mycobacterium tuberculosis* complex. \*Medical harmfulness domain evaluation rubric (score): severely harmful (1), moderately harmful (2), mildly harmful (3), minimally harmful (4), harmless (5)

The running cost of GenAI chatbots have reduced substantially over time. At the time of testing, GPT-4.0’s operating costs were £0·0474 per 1,000 tokens for input and £0·0948 per 1,000 tokens for output, with average costs for scenario input and output calculated to be £0·0204 and £0·0408, respectively. Within the subsequent six months, the average cost per 1,000 tokens for input and output decreased by approximately 50% for GPT-4.0 while costs for Claude 2 remained unchanged. Notably, Gemini Pro has transitioned to a free service model. Currently, the operating costs for frontier models, such as: GPT-4o, GPT-4 Turbo, Claude 3 Opus, and Gemini 1.5 Pro are comparable. As competition among GenAI models intensify, the cost disparity between proprietary models (GPT-4.0, Gemini 1.5 Pro, Claude 3) and open-source models (Llama 3, Meta Platforms, Inc., California, U.S.; Mistral 7B, Mistral AI, Paris, France) is expected to narrow. This market trend will enable healthcare institutions to integrate state-of-the-art AI technologies into their clinical workflow at a cost-effective manner.

### Limitations

Several limitations are identified in this study. First, the research was conducted at a tertiary/quaternary referral centre, where the case mix may not be representative of the broader healthcare system in HK, therefore limiting the generalisability of our findings.

Second, for fair comparisons, standardised, complete, and verified data were used to create case scenarios. However, the level of clinical detail and available patient data in these scenarios may not fully encapsulate the variability and nuances of real-life hospital settings. Since AI system performance is highly dependent on the quality of input data, it is important to recognise that AI-generated responses may be constrained in actual clinical practice.

Third, our study did not incorporate domain-specific healthcare AI models, such as Med-PaLM 2^26^ or MEDITRON^27^, which are designed to enhance performance through specialised pre-training, fine-tuning, and advanced prompt engineering. As AI technology continues to advance rapidly, these models are expected to achieve clinical safety and reliability shortly. It is important for stakeholders to stay informed about the latest developments to fully leverage AI’s potential in healthcare.

The authors emphasis that AI systems should not replace human clinicians or their judgements. Instead, future research should prioritise comparative analyses between traditional clinical care and AI-enhanced healthcare delivery to unlock the full potential of AI technologies across diverse healthcare settings. From a patient engagement perspective, multimodal capabilities of AI systems can significantly enhance doctor-patient communication, aiding in the explanation of complex medical concepts through multimedia channels, thereby empowering patient, reinforcing their autonomy, and fostering better shared decision-making.^28^ In terms of cross-specialty collaboration, AI could efficiently capture the entirety of the patient’s clinical journey across the full spectrum of the healthcare ecosystem—primary, secondary, tertiary, and community care.^29^ Integration of unstructured health data into the chronological profile of the patient could enable powerful insights into health state, thereby facilitating timely and proactive health interventions. Additionally, real- time monitoring of communicable diseases and available healthcare resources [e.g., personal protective equipment (PPE), vaccines, treatments, laboratory reagents…] should be guided by big data and analysed by AI, allowing precise and equitable distribution of resources and effective management of supply chain constrains, thereby enabling rapid public health interventions.^30^

## Data Availability

All relevant data are within the manuscript and its Supporting Information files. (The full database of scores will be available in subsequent submissions.)

## Contributors

E.K.Y.C conceptualized the study, curated the data, led the investigation, conducted formal analysis, designed the methodology, developed the software, created visualizations, and was primarily responsible for writing the original draft as well as reviewing and editing the manuscript. S.S contributed through supervision of the investigation and by participating in the manuscript review and editing process. S.S.Y.W, A.R.T, M.H.C, K.H.Y.C, A.W.T.L, and W.C.W were involved in conducting the investigation. M.Y.Z.N was responsible for data curation. K.Y.Y was involved in reviewing and editing the manuscript. T.W.H.C, as the corresponding author, took on roles in conceptualization, data curation, investigation, formal analysis, project administration, methodology design, supervision, validation, visualization, and writing both the original draft and the review & editing of the manuscript. All authors had full access to all the data in the study and had final responsibility for the decision to submit for publication. Both E.K.Y.C and T.W.H.C verified the data and contributed equally to the study.

## Funding

There was no funding source for this study.

## Notes

### Competing Interest Statement

The authors have declared no competing interest.

### Clinical Protocols

https://www.medrxiv.org/content/10.1101/2024.03.01.24303593v1

### Funding Statement

The author(s) received no specific funding for this work.

### Author Declarations

This study was approved by the University of Hong Kong and Hospital Authority Hong Kong West Cluster Institutional Review Board (UW 24-108).

## References

1. Denniston AK, Liu X. Responsible and evidence-based AI: 5 years on. The Lancet Digital Health 2024;6(**5**):e305–e7.

2. Li H, Moon JT, Purkayastha S, Celi LA, Trivedi H, Gichoya JW. Ethics of large language models in medicine and medical research. The Lancet Digital Health 2023;5(**6**):e333–e5.

3. Howard A, Hope W, Gerada A. ChatGPT and antimicrobial advice: the end of the consulting infection doctor? The Lancet Infectious Diseases 2023;23(**4**):405–6.

4. Schwartz IS, Link KE, Daneshjou R, Cortés-Penfield N. Black box warning: large language models and the future of infectious diseases consultation. Clinical Infectious Diseases 2024;78(**4**):860–6.

5. Sarink MJ, Bakker IL, Anas AA, Yusuf E. A study on the performance of ChatGPT in infectious diseases clinical consultation. Clinical Microbiology and Infection 2023;29(**8**):1088–9.

6. Armitage R. Large language models must serve clinicians, not the reverse. The Lancet Infectious Diseases 2024.

7. Langford BJ, Branch-Elliman W, Nori P, Marra AR, Bearman G, editors. Confronting the Disruption of the Infectious Diseases Workforce by Artificial Intelligence: What This Means for Us and What We Can Do About It. Open Forum Infectious Diseases; 2024: Oxford University Press US.

8. Chiu KYE, Chung TW-H. Protocol For Human Evaluation of Artificial Intelligence Chatbots in Clinical Consultations. medRxiv 2024:2024.03.01.24303593.

9. Best practices for prompt engineering with OpenAI API: OpenAI; 2024 [Available from: https://help.openai.com/en/articles/6654000-best-practices-for-prompt-engineering-with-openai-api. (accessed 12 January 2024).

10. The Art of AI Prompt Crafting: A Comprehensive Guide for Enthusiasts: OpenAI; 2023 [Available from: https://community.openai.com/t/the-art-of-ai-prompt-crafting-a-comprehensive-guide-for-enthusiasts/495144. (accessed 12 January 2024).

11. Prompt engineering: OpenAI; 2023 [Available from: https://platform.openai.com/docs/guides/prompt-engineering. (accessed 12 January 2024).

12. Wang L, Chen X, Deng X, Wen H, You M, Liu W, et al. Prompt engineering in consistency and reliability with the evidence-based guideline for LLMs. npj Digital Medicine 2024;7(1):41.

13. Prompt engineering techniques: Microsoft Corporation; 2023 [Available from: https://learn.microsoft.com/en-us/azure/ai-services/openai/concepts/advanced-prompt-engineering?pivots=programming-language-chat-completions. (accessed 12 January 2024).

14. Retrieval Augmented Generation (RAG) and Semantic Search for GPTs: OpenAI; 2024 [Available from: https://help.openai.com/en/articles/8868588-retrieval-augmented-generation-rag-and-semantic-search-for-gpts. (accessed 31 May 2024).

15. Török E, Moran E, Cooke F. Oxford handbook of infectious diseases and microbiology. 2nd ed: Oxford University Press; 2016.

16. Mitchell RN, Kumar V, Abbas AK, Aster JC. Pocket Companion to Robbins & Cotran Pathologic Basis of Disease E-Book. 9th ed: Elsevier Health Sciences; 2016.

17. Sabatine MS. Pocket medicine (Pocket notebook series). 8th ed: Wolters Kluwer Health; 2022.

18. Gilbert DN, Chambers HF, Saag MS, Pavia AT, Boucher HW. The Sanford guide to antimicrobial therapy 2022. Antimicrobial Therapy 2022.

19. Hinton G, Vinyals O, Dean J. Distilling the knowledge in a neural network. arXiv preprint arXiv:150302531 2015.

20. Tang L, Sun Z, Idnay B, Nestor JG, Soroush A, Elias PA, et al. Evaluating large language models on medical evidence summarization. NPJ Digit Med 2023;6(**1**):158.

21. Von Elm E, Altman DG, Egger M, Pocock SJ, Gøtzsche PC, Vandenbroucke JP. The Strengthening the Reporting of Observational Studies in Epidemiology (STROBE) statement: guidelines for reporting observational studies. The lancet 2007;370(9596):1453–7.

22. Goodman RS, Patrinely JR, Stone CA, Zimmerman E, Donald RR, Chang SS, et al. Accuracy and reliability of chatbot responses to physician questions. JAMA network open 2023;6(**10**):e2336483-e.

23. OpenAI Language Models Pricing: OpenAI; 2024 [Available from: https://openai.com/api/pricing/. (accessed 12 April 2024).

24. Claude API: Anthropic PBC; 2024 [Available from: https://www.anthropic.com/api. (accessed 12 April 2024).

25. Gemini API Pricing: Google LLC; 2024 [Available from: https://ai.google.dev/pricing. (accessed 12 April 2024).

26. Singhal K, Tu T, Gottweis J, Sayres R, Wulczyn E, Hou L, et al. Towards expert-level medical question answering with large language models. arXiv preprint arXiv:230509617 2023.

27. Chen Z, Cano AH, Romanou A, Bonnet A, Matoba K, Salvi F, et al. Meditron-70b: Scaling medical pretraining for large language models. arXiv preprint arXiv:231116079 2023.

28. Qiu J, Yuan W, Lam K. The application of multimodal large language models in medicine. The Lancet Regional Health–Western Pacific 2024;45.

29. Patel SB, Lam K. ChatGPT: the future of discharge summaries? The Lancet Digital Health 2023;5(**3**):e107–e8.

30. Feng J, Phillips RV, Malenica I, Bishara A, Hubbard AE, Celi LA, et al. Clinical artificial intelligence quality improvement: towards continual monitoring and updating of AI algorithms in healthcare. npj Digital Medicine 2022;5(**1**):66.

